# Local REM sleep-N1-wake sleep stage mixing in narcolepsy type 1

**DOI:** 10.64898/2026.02.14.26346110

**Authors:** Constantin Weberpals, Adrien Specht, Natasja Bonde Andersen, Mads Olsen, Yves Dauvilliers, Giuseppe Plazzi, Lucie Barateau, Fabio Pizza, Francesco Biscarini, Jing Zhang, Han Yan, Ambra Stefani, Birgit Hogl, Matteo Cesari, Seung Chul Hong, Dmitri Volfson, Poul Jennum, Andreas Brink-Kjaer, Emmanuel Mignot

## Abstract

Type 1 narcolepsy (NT1), a disorder caused by the loss of hypocretin/orexin transmission, is characterized by daytime sleepiness and symptoms where Rapid Eye Movement (REM) sleep, a state normally occurring from middle to late in the night, can intermingle with wakefulness. This results in cataplexy and sleep paralysis, episodes of muscle paralysis when awake, or in the generation of dream-like hallucinations and vivid dreaming, periods of visual imagery or sensory experiences that occur while awake, notably when falling asleep (hypnagogic hallucinations) or lingering dreams with over-realistic recall. Using deep learning of nocturnal sleep polysomnography (PSG) signals (EEG, EMG and EOG) applied to sleep stage scoring, we found that NT1 shows abnormally short wake to REM sleep transitions and occurrences of abnormal sleep stages probabilities of wake, REM sleep and N1 (very light NREM) sleep abnormally co-occurs (sleep stage mixing). Interestingly, although presence of these during sleep enables NT1 diagnosis with performances similar to gold standard diagnostic procedure, the multiple sleep latency test (MSLT), the cortical localization of these dissociations remains unclear. In this work, we used electrode specific predictions of sleep stages to explore if these are global or observed at the local cortical level. Surprisingly, although sleep stage mixing was preeminent between REM sleep, N1 and wake across all electrodes, it was found to fluctuate across locations, with stronger fluctuations found in frontal and central locations, notably in the dominant (left) hemisphere. The strongest single discriminator for NT1 was N1-REM stage mixing across central electrodes (C3-C4), showing 4.3-fold higher dissociation in NT1 patients (Cohen’s d = 0.61). Analysis of sleep stage dissociations across varying time scales revealed that windows lasting several minutes were most predictive of NT1 status, aligning with the duration of clinically reported symptoms of dissociated REM sleep in narcolepsy. Local N1-W-REM sleep dissociations correlated with CSF orexin/hypocretin levels and severity as measured using MSLT. The predominance of stage mixing in frontal and central regions, areas typically associated with executive and motor control, may contribute to the partial preservation of awareness during dissociated REM phenomena. Further, self-reports of hypnagogic hallucinations correlated best with dissociations involving occipital locations, in agreement with its usual visual content. Coherence analysis was also conducted but did not reveal additional insight. These results suggest that orexin deficiency destabilizes REM sleep organization across cortical projection area contributing both to REM sleep dissociation and to abnormal state transitions observed in NT1.

## Introduction

Sleep is composed of two main states, Rapid Eye Movement (REM) Sleep and Non-REM (NREM) sleep, sleep states where EEG activity is generated by a complex dialogue between the thalamus, the hippocampus and cortical region^1,2^. NREM sleep is often separated into stage 1, a transitory stage between wake and sleep where occipital alpha activity (present when eyes are closed and the subject awake) disappears and electroencephalogram (EEG) slows down in frequency; stage 2, characterized by sleep spindles and the appearance of K-complexes (lasting a few seconds); and stage 3, dominated by slow wave activity. Sleep onset, the associated loss of awareness that accompany sleep, normally occurs in stage 1 or stage 2^3,4^.

NREM sleep is regulated by cortical thalamic interactions^5^ and reflects, at least partially, prior usage during wake at the local cortical circuitry level. As an example, prior experiences stimulating subareas of the motor cortex during the day are associated with rebound NREM sleep, as measured by slow wave sleep activity, in the corresponding area during the following night^6,7^. A circadian influence is also important for NREM regulation and helps maintaining wake in the evening, when sleep debt is highest as it has accumulated all day^8^. Synaptic reorganization, changes in brain fluid dynamics and many other phenomena occur during NREM sleep^9,10^, most notably synaptic downscaling. NREM sleep, and notably spindles, are important for memory consolidation^8,11^, and are likely energy-saving for the organism^12^. Although some degree of cognition occurs in NREM sleep, dream recall from NREM is poor, and more so in stage 3 sleep^13^.

In contrast, REM sleep is characterized by fast, low-amplitude EEG with vivid dream recall, muscle atonia, and rapid eye movements. It alternates between tonic REM sleep, marked by sustained atonia and few or no eye movements, and phasic REM sleep, characterized by bursts of rapid eye movements and occasional muscle twitches. In normal sleepers, REM sleep typically occurs 90-120 min after sleep onset in the evening and recurs by cycles interspersed by NREM sleep to become more preeminent in the morning hours when the circadian drive for REM sleep is strongest. REM sleep represents 20% of total sleep time in adults, showing a steep decline from infancy to adolescence and a more gradual decrease with aging^14^. It is independently homeostatically regulated, with strong gating by the central circadian clock^15–18,12^.

Genesis of REM sleep arises from the pons^19^, with ascending projections regulating cortical activation and eye movements, while descending projections produce atonia through inhibition of the monosynaptic spinal reflex. Ponto-geniculo-occipital (PGO) waves for example, progress from the pons to the thalamus and occipital cortex, likely supporting REMs and associated imagery^20^, although in humans, presence and nature of PGOs are controversial^21–23^. REM sleep as observed in the cortex is generally considered as a global phenomenon, although recent work indicates central and frontal dominance of theta rhythms and sawtooth waves during REM sleep^24,25^. Importantly however, heterogeneity and rapid switching of saw tooth waves from one region to the next, a combination of slow and theta activity characteristic of REM sleep occurs, perhaps correlating with the complex representation of dreams during REM sleep^26^. Clearly, although REM sleep is often simplified as the result of homogeneous ascending activation, aspects of REM sleep manifest differentially in subcortical regions. Recent intracranial EEG studies have demonstrated that even NREM-to-REM sleep transitions show regional heterogeneity, with transitions occurring first in lateral occipital cortex and preceding scalp transitions by nearly 2 minutes, while frontal regions show the latest transitions^27^.

Following REM sleep, remembrance of dreams is generally fleeting^28^, with most individuals never remembering dreaming or being paralyzed. An exception to this occurs in type 1 narcolepsy^29,30^, a disorder caused by a lack of orexin^31^, a wake promoting neuropeptide that also strongly inhibits REM sleep^32^. Orexin is a generally excitatory transmitter, and orexin neurons project widely throughout the entire neuroaxis, including on adrenergic, serotoninergic and histaminergic neurons known to be wake active and silent during REM sleep^33,34^. Projections to cholinergic neurons of the basal forebrain and to the cortex, layer six b, are also notable^33^. In NT1 patients, dissociations of REM sleep and wake are common and severe, so that patients can be paralyzed but awake at sleep onset, while napping or during the night, a symptom called sleep paralysis^29,35^. These patients also have vivid dream like experiences while still conscious, often at sleep onset or when very sleepy, or in the middle of the night^29^. These “half dream” hallucinatory experiences are frequently visual and leave a strong memory imprint that can make the sleeper feel as it truly happened^36^.

In patients with type 1 narcolepsy, we recently found that sleep is not only disturbed in its sleep stage sequencing (for example short REM sleep latency after sleep onset)^37,38^ but also presents with true intra sleep stage dissociations^39^. During these periods, features of REM sleep, wake and stage 1 are intermingled at frequencies higher than those describing microarchitecture features (1-3 sec), a speed like what is needed to compose a complex thought^39^. As a result, NT1 patients are not only showing premature REM sleep early in the night (sleep onset REM periods) or abnormal wake to REM sleep transition during night sleep, but also genuine periods of mixed states, perhaps sustaining symptomatic experiences of vivid dreaming, hypnagogic hallucinations or sleep paralysis^39,40^. These findings replicated in humans the evidence of labile boundaries between sleep and wake described in narcoleptic mice^41^.

Interestingly, these dissociations are conveniently detected by sleep staging neural networks, which, in many NT1 patients, will show sleep periods where the predicted probability of REM sleep, wake and N1 is intermingled without a clear dominant state, a rare occurrence in controls^39^. This phenomenon is best visualized through hypnodensity plots, which display the continuous probability distribution of sleep stages across time rather than discrete labels. In fact, a feature combination of abnormalities in sleep stage sequencing and hypnodensities (sleep stage probability distribution per epoch) has sensitivity and specificity for diagnosing NT1 above 95%^39,42^. This mirrors performance of the current gold standard, the Multiple Sleep Latency (MSLT) test, a test conducted during the daytime where naps are scheduled, sleep latency measured and the occurrence of REM sleep in these naps noted^37^. In this test, a mean sleep latency below 8 minutes and occurrence of ≥ 2 SOREMPs in 4-5 naps plus nocturnal PSG is diagnostic for NT1.

In this study, we aimed to determine whether sleep stage mixtures in NT1 result from localized cortical differences or global mixing across the entire neuroaxis. We further tested whether such localized effects shift rapidly between cortical regions or remain stable within specific areas over longer periods. To do so, we extended our probabilistic framework using a specialized neural network trained on healthy subjects to perform sleep stage scoring separately for different modalities (EEG, EOG, and EMG) and cortical regions (occipital, central and frontal). We also conducted coherence analyses to examine frequency-band desynchronization. Together, these approaches were designed to assess whether local sleep stage dissociations could account for the heterogeneous expression of REM-related phenomena such as sleep paralysis, hypnagogic hallucinations, and conscious dreaming in NT1, and to explore their potential as novel diagnostic markers.

## RESULTS

### Regional REM sleep Dissociations Distinguishes NT1 from Controls

Differences in sleep stage synchronization may be better assessed by comparing sleep stage mixing, a strong feature of narcolepsy^39,40^. To investigate whether sleep stage dissociations occur at the regional level in narcolepsy type 1 (NT1), we used a U-sleep based neural network to generate channel-specific hypnodensity distributions and evaluated area of stage uncertainty (mixed stage) at each electrode location rather than within a single global sleep stage classification^43^. Sleep stage dissociations were defined similarly to Stephensen et al.^39^ and Olsen et al^40^, and regional differences in dissociations identified as absolute differences in identified mixed states across electrode locations (see Figure 1).

**Fig 1.**
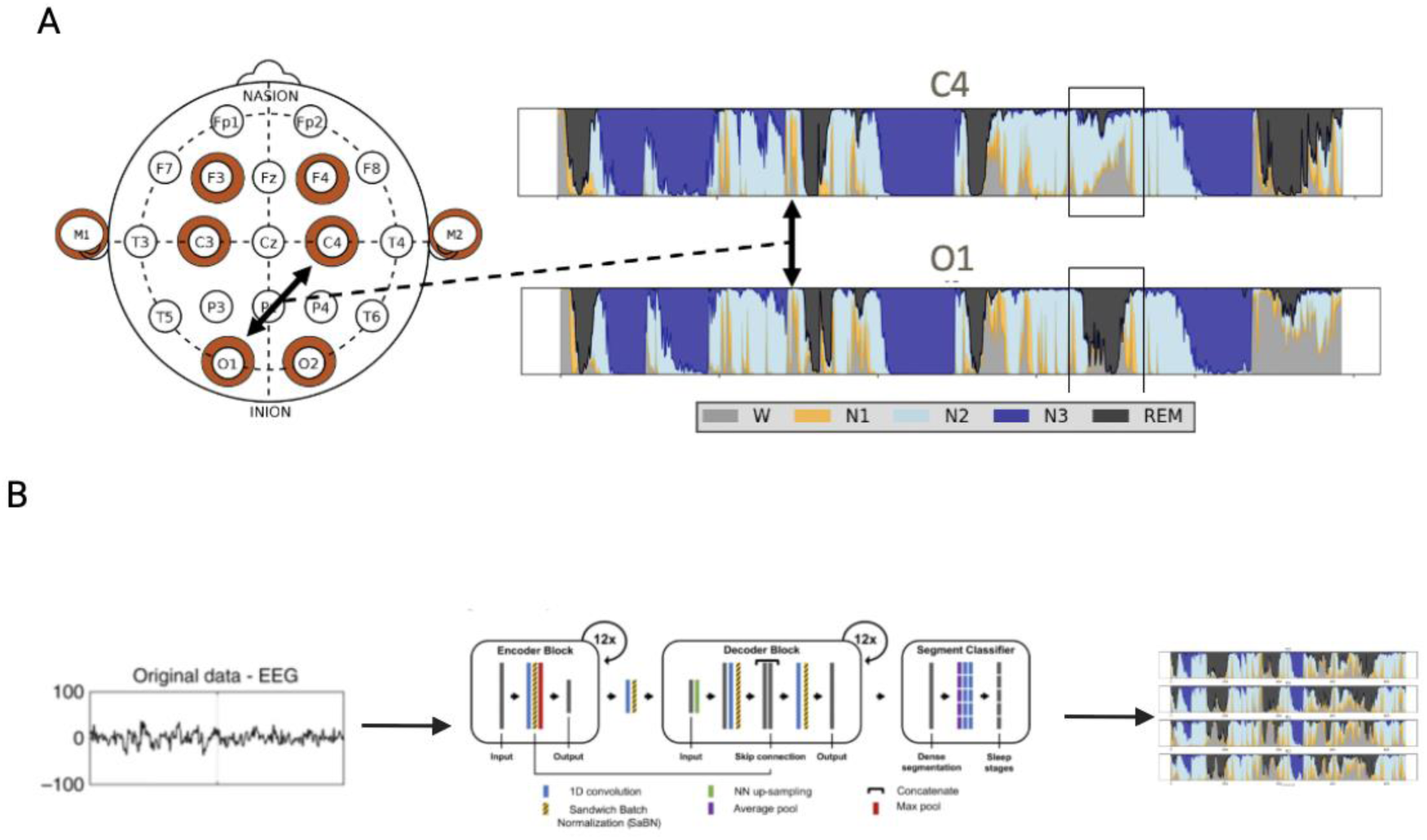
Methodology for Detecting Regional Sleep Stage Dissociations in Narcolepsy Type 1. A) Conceptual overview of regional sleep stage analysis. The 10-20 electrode system (left) shows key recording positions, with highlighted electrodes (C3, C4, O1, O2) used for regional analysis. Hypnodensity plots for central (C4) and occipital (O1) channels demonstrate how different brain regions can simultaneously exhibit distinct sleep stage probability distributions. Sleep stages are color-coded: Wake (gray), N1 (orange), N2 (light blue), N3 (dark blue), and REM (black). B) U-Sleep neural network architecture^43^ for channel-specific sleep staging. Raw EEG signals are processed through sequential encoder-decoder blocks with skip connections to generate probabilistic sleep stage distributions for each electrode independently. The network outputs continuous hypnodensity graphs representing probability distributions across all sleep stages for each analyzed channel.

As expected, NT1 patients display mixed states, notably across N1, wake and REM sleep when evaluated globally using hypnodensity with all electrodes available (see supplementary table 2). Additionally, however, NT1 patients also showed significantly greater dissociation differences between brain regions compared to healthy controls. This was found across multiple similarity measures and temporal resolutions (1s, 3s, 15s and 30s epoch). Analysis of the most significant features (p < 0.001) revealed 44 total dissociation differences across electrode pairs, with clear regional organization (Figure 2). Most notably, the C3-C4 central electrode pair showed dominance, accounting for 18 of 44 highly most significant features (40.1%, all p < 0.001), more than any other electrode combination (Figure 2B). The strongest single discriminator was a difference in N1-REM stage mixing (Cohen’s d = 0.61, OR = 4.26, p < 0.001) across C3-C4, where NT1 patients showed 4.3-fold higher dissociation when compared to controls (NT1: 4.602 ± 1.676 vs. Controls: 2.829 ± 1.123). Not surprisingly, as reported globally, the most significant regionally different dissociation features involved REM sleep, highlighting the importance of this stage in the pathophysiology of NT1 (Figure 2A).

**Figure 2.**
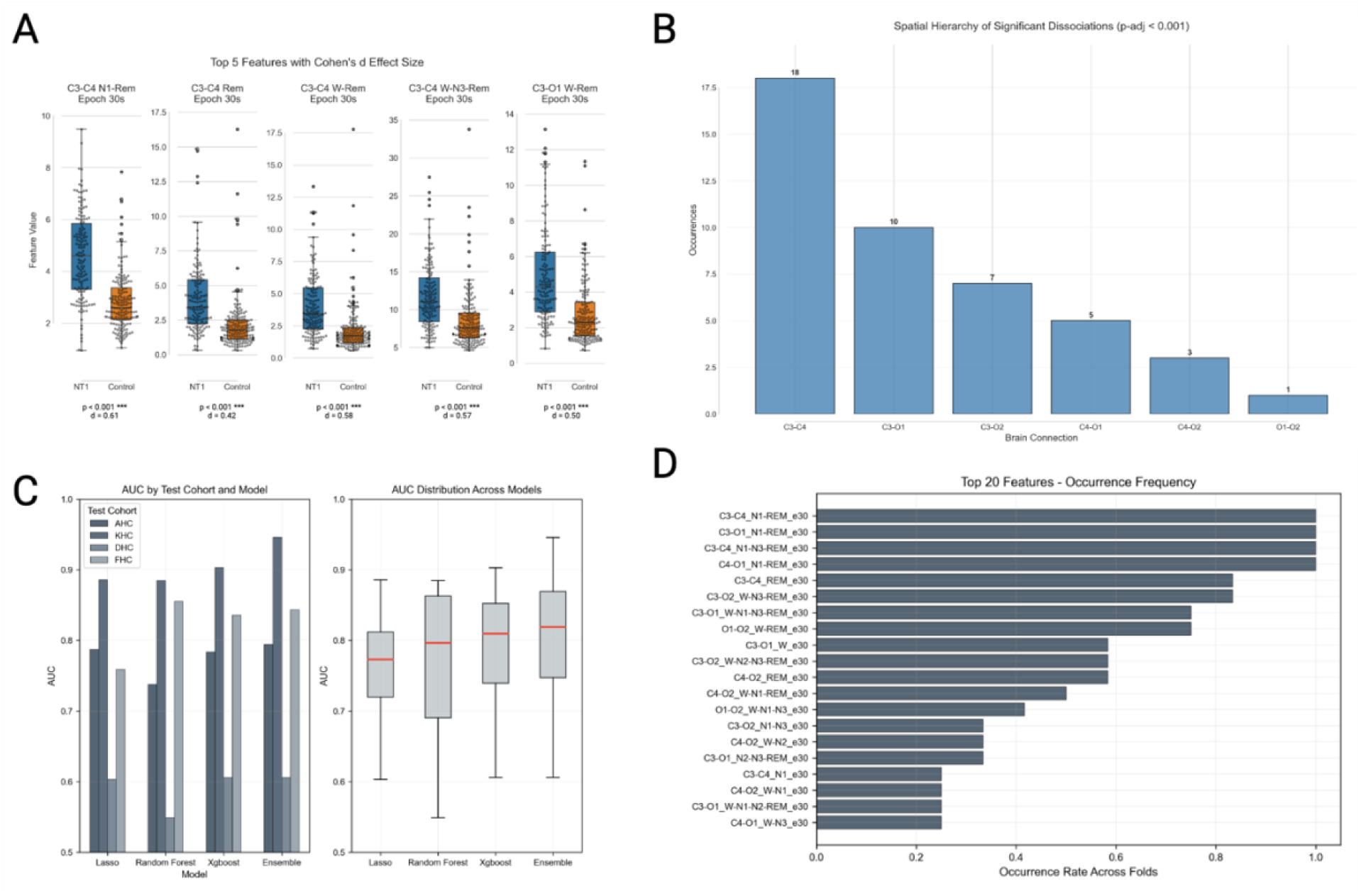
Regional EEG Sleep Stage Dissociations in Narcolepsy Type 1: Hierarchical Spatial Organization and Diagnostic Utility. A) Comparing top stage mixing dissociation features between NT1 patients (blue) and controls (orange) across different electrode pairs. All features show significantly higher dissociation in NT1 (p < 0.001). B) Frequency distribution of significant features (p < 0.001) showing hierarchical spatial organization. C) Leave-one-cohort-out cross-validation performance across four clinical sites demonstrates robust diagnostic generalizability. Bar plots show ensemble and individual algorithm performance across cohorts (LASSO, Random Forest, XGBoost) while box plots show mean AUC values by algorithm. D) Feature importance analysis based on selection frequency across algorithms reveal N1-REM as consistent pattern and highlights dominance of REM-based features. Frontal features are not included here as the could only be assessed on a smaller sample size.

### Regional Analysis of Stage Mixing Reveals Clear Hierarchical Organization

The most significant feature showed anatomical specificity (Figure 2B): C3-C4 central pair (18 features) >> C3-O1 left centro-posterior (13 features) > C3-O2 cross-hemispheric (10 features) > C4-O1 cross-hemispheric (7 features) > C4-O2 right centro-posterior (4 features) >> O1-O2 occipital pair (1 feature). This 18:1 ratio between central and occipital electrode pairs suggests dissociations were strongest in sensorimotor regions. Central-occipital derivations showed intermediate vulnerability, with C3-O1 demonstrating 3.25 times more significant features than C4-O2 (13 vs 4), revealing a trend toward left-hemisphere predominance.

In the subset of subjects with frontal electrode coverage (N=40 NT1, N=39 controls), frontal-central dissociations demonstrated strong effects with high odds ratios, suggesting a frontal to occipital gradient of dissociation. The C4-F3 electrode pair showed significant dissociations like N1-REM stage mixing (Cohen’s d = 0.77, OR = 6.21, p < 0.01) W-N1-REM mixing (Cohen’s d = 0.65, OR = 8.99, p < 0.05). Contralateral connections showed higher odds ratios (mean OR = 6.48 ± 1.24) compared to ipsilateral pairs (mean OR = 3.61 ± 1.72). This pattern was consistent across both the large central-occipital cohort and the smaller frontal electrode subset.

### Multi-Cohort Validation of Regional Dissociation Features for NT1 Diagnosis

To evaluate diagnostic robustness and generalizability, we used a leave-one-cohort-out (LOCO) cross-validation methodology using our multi-center dataset. This approach tests the most clinically relevant scenario, whether models trained with data obtained at one sleep center can diagnose NT1 at previously unseen clinical sites. LASSO regression (L1-regularized logistic regression), Random Forest, XGBoost, and an ensemble method combining all three algorithms were used. The ensemble approach demonstrated superior overall performance (AUC = 0.797 ± 0.123, AP = 0.787 ± 0.158), outperforming individual algorithms while maintaining robust generalization across sites. XGBoost emerged as the strongest individual algorithm (AUC = 0.782 ± 0.110, AP = 0.777 ± 0.158), followed closely by LASSO (AUC = 0.759 ± 0.101) and Random Forest (AUC = 0.757 ± 0.132) as displayed in Figure 2C. These results indicate that regional differences in sleep stage mixing are robust across cohorts and have diagnostic value.

### Site-Specific Performance Reveals Clinical Heterogeneity and Robustness

LOCO validation revealed between-site variability, with test cohort AUC ranging from 0.606 (DHC) to 0.946 (KHC). Three of four cohorts demonstrated stronger diagnostic performance (KHC: AUC = 0.946, FHC: AUC = 0.844, AHC: AUC = 0.794), with the Danish cohort showing more modest results (DHC: AUC = 0.606). The KHC cohort demonstrated exceptional performance across all algorithms (ensemble AUC = 0.946, individual algorithms all >0.88). The FHC cohort showed balanced strong performance (ensemble AUC = 0.844), with Random Forest performing particularly well (AUC = 0.855). The AHC cohort maintained good diagnostic accuracy (ensemble AUC = 0.794) with consistent performance across algorithms. The DHC cohort presented the greatest challenge (ensemble AUC = 0.606), showing modest performance across all methods. Importantly, even this most challenging cohort achieved above-chance classification, indicating that regional dissociation patterns are detectable across diverse clinical settings, though with varying sensitivity.

### C3-C4 N1-REM Dissociation Emerges as Consistent Biological Pattern Across Sites and Algorithms

In all cases regional differences in C3-C4 N1-REM stage mixing emerged as the most important feature across all algorithms and cohorts, ranking first in LASSO (coefficient = 1.748 ± 1.023, appearing in 4/4 cohorts, Figure 2D), Random Forest (importance = 0.106 ± 0.057), and consistently high in XGBoost analyses. This remarkable consistency across different analytical approaches and clinical sites validates C3-C4 N1-REM dissociation as a core pathophysiological signature of NT1.

LASSO regression showed preference for high-magnitude dissociation features, with larger coefficients for complex multi-stage mixing patterns (W-N1-REM, N1-N3-REM combinations). Random Forest emphasized broad N1-REM patterns across multiple electrode pairs, suggesting ensemble benefits from diverse regional perspectives on the same pathophysiological process. XGBoost identified more nuanced patterns, including complex multi-stage features that complement the simpler N1-REM patterns. Feature stability across cross-validation folds was strong, with C3-C4 N1-REM appearing in 100% of training models and maintaining top rankings across sites. The mean coefficient for C3-C4 N1-REM in LASSO models (1.748 ± 1.023) was more than 40% larger than the second-ranked feature, emphasizing its dominant discriminative power.

Frontal locations showed larger effect sizes but, as these could only explored in a smaller subset, were less statistically significant. Importantly, frontal EEG is frequently contaminated by EOG activity, and some of the effects observed here could derived from EOG differences, which were very preeminent in these datasets (see below), although this is not likely as fronto-occipital pairs showed less differences than frontocentral ones. Additional studies including higher density electrode set up will be needed to pursue these findings at higher spatial resolution.

### Regional Dissociations Reflect Spatial Sleep State Mixing Beyond Increased Transitions

To determine whether regional sleep stage dissociations simply reflect increased sleep stage transitions in NT1, we quantified transition frequency at each electrode. NT1 patients showed significantly higher transition counts across most electrodes (C3, C4, O1, O2, F3: all p<0.001), with transition frequencies highly correlated across electrodes in both NT1 (r=0.76-0.92) and controls (r=0.72-0.93) (Supplementary Figure 3). However, when transition count was included as a covariate, regional dissociation features remained highly significant after adjustment, with other covariates like age (p=5.12×10⁻⁶) and cohort (p=3.62×10⁻¹¹) being substantially more influential factors than transition count (p=6.62×10⁻³).

### Optimal Resolution and Sustained Dissociation Patterns

To determine optimal temporal resolution of regional dissociations, we analyzed variance ratios across multiple epoch lengths (1s, 3s, 15s, 30s) for all significant electrode pairs and features. Signal-to-noise ratios showed consistent improvement with longer epoch durations, with most features reaching optimal discrimination at a resolution between 15-30 seconds (Figure 3A). This temporal optimization suggests that regional sleep stage dissociations in NT1 operate at behaviorally relevant timescales (15-30s), consistent with perceived symptomatology such as abnormal dreaming or sleep paralysis rather than artifact-driven rapid fluctuations.

**Figure 3.**
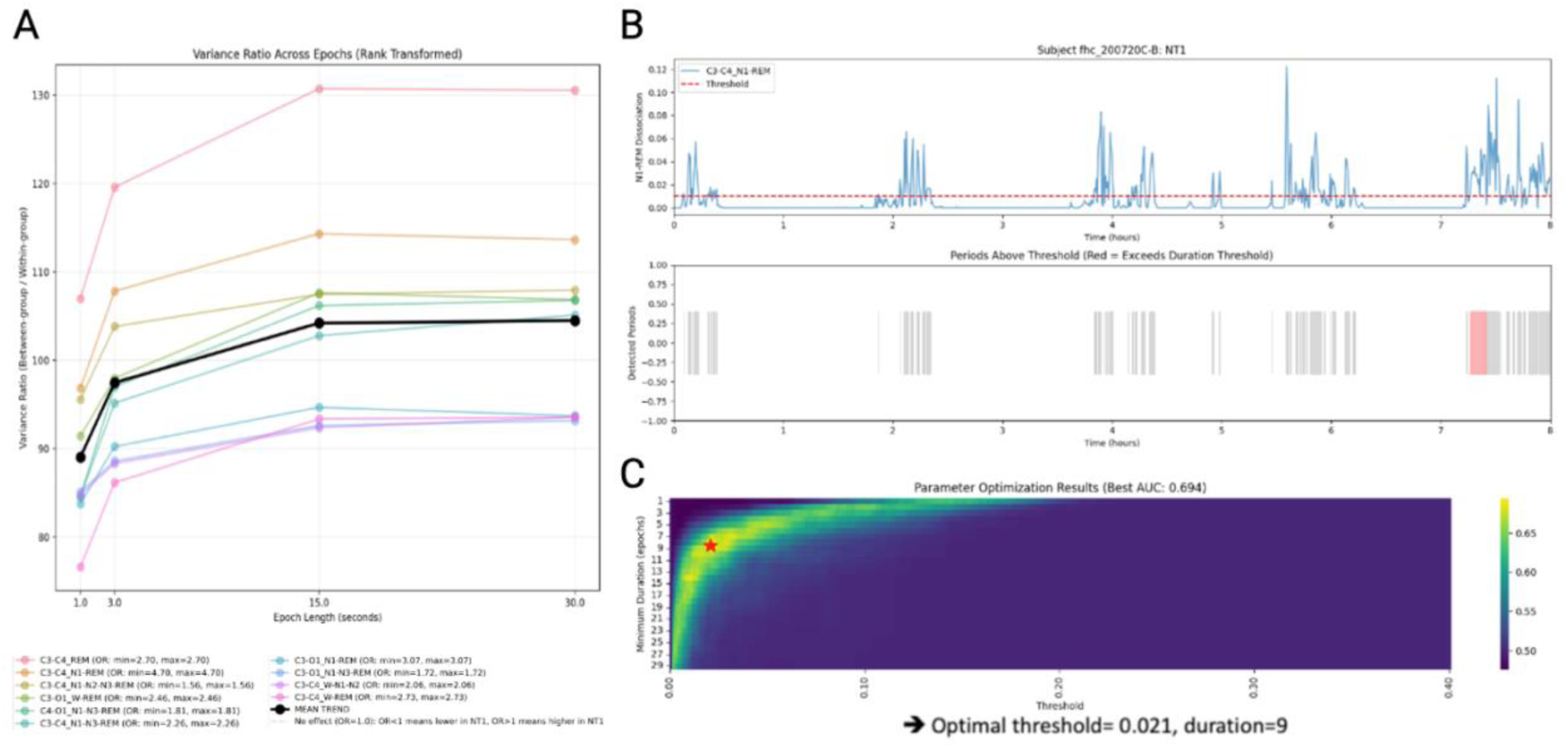
Temporal dynamics of sleep stage dissociation in narcolepsy. A) Variance ratios (between-group variance divided by within-group variance) are shown for key electrode pair and stage-mixing combinations across epoch lengths from 1 to 30 seconds. B) Time course of N1-REM dissociation in a NT1 patient over an 8-hour sleep recording. The upper plot shows C3-C4 dissociation magnitude with detection threshold (red dashed line), while the lower plot identifies periods exceeding both magnitude and duration thresholds (red highlighted region). C) Parameter optimization heatmap showing diagnostic performance (AUC) across different dissociation thresholds and minimum duration requirements. Optimal parameters (red star) were identified at threshold=0.021 and duration=9 epochs of 15 seconds (2.25 minutes), yielding maximum AUC of 0.694 for distinguishing NT1 from controls.

Beyond instantaneous dissociation magnitudes, we investigated whether prolonged episodes of regional disconnection provided additional diagnostic information. Sustained dissociation episodes were defined as continuous periods where dissociation magnitude exceeded an optimized threshold (see Figure 3C and Methods). Analysis of sustained C3-C4 N1-REM dissociation periods (which was previously detected as the top feature) revealed that NT1 patients experience significantly longer episodes of regional sleep stage mixing compared to controls (Fig. 2B). The moderate diagnostic performance of sustained episodes (AUC = 0.694) compared to mean dissociation features (AUC > 0.80 on combined dataset) indicates that while prolonged disconnections are characteristic of NT1, the overall magnitude and frequency of dissociations is more diagnostic, revealing that regional dissociations in NT1 are not transient artifacts but represent sustained periods of regional sleep-stage dissociations.

### Dissociation Magnitude Rather Than Direction Distinguishes NT1

Analysis of directional dissociations (which electrode showed higher stage probability) revealed expected physiological gradients—for example, frontal > central > occipital N3 probability (Supplementary Figure 4)—but showed low discriminative power between NT1 and controls when aggregated across the night. NT1 patients showed regional sleep architecture abnormalities that evolved over time: elevated REM sleep probability in frontal and central regions during early night quarters (Q1-Q2), persistently increased N1 probability across most locations throughout the night, and reduced N2 probability particularly in the second half of the night (Supplementary Figure 6). However, despite these clear directional patterns, they showed low discriminative power between NT1 and controls when aggregated across the night compared to the absolute dissociation features. This indicates that the pathology lies in the fluctuating magnitude of regional dissociations rather than consistent directional biases, supporting the interpretation that NT1 is characterized by temporal instability in sleep state synchronization across cortical regions rather than fixed regional sleep stage abnormalities.

### System-wide Dissociation Patterns

To investigate whether regional sleep stage dissociations extend beyond cortical networks, we next analyzed cross-modal dissociations across EEG electrodes and EMG (muscle) and EOG (eye movement) channels. Supplementary Figure 5 illustrates exemplary hypnodensity profiles across all modalities (EEG electrodes, EMG, and EOG) comparing a healthy control with synchronized sleep stage probabilities across channels to an NT1 patient exhibiting marked dissociations, where sleep stage probabilities diverge across cortical regions and physiological systems within the same time periods For this analysis we only utilized our complete multi-cohort dataset across 5 cohorts (N=452 NT1 patients and N=271 controls), as all recording sites included standard EMG and EOG channels, providing greater statistical power than the regional EEG analysis.

#### EEG-EMG Cortico-Motor tone Disconnections

EEG-EMG dissociations showed more moderate effect compared to oculomotor patterns. The strongest discriminator was W-REM dissociation (Cohen’s d = 0.52, OR = 2.49, p < 0.001), with other highly significant features such as W-N1-REM (Cohen’s d = 0.42, OR = 2.47, p < 0.001) (Fig. 4B). NT1 patients showed 2-2.5-fold higher odds of exhibiting elevated EEG-EMG dissociation compared to controls. For W-N1-REM mixing, NT1 patients demonstrated mean values of 33.320 ± 2.715 compared to 31.091 ± 2.020 in controls. Interestingly, selected EEG-EMG features showed negative effects (such as N2 stage mixing with Cohen’s d = −0.46), suggesting compensatory mechanisms. The pattern of EEG-EMG dissociations directly relates to sleep paralysis pathophysiology, where REM atonia mechanisms persist inappropriately during sleep-wake transitions, causing patients to experience conscious awareness while remaining physically paralyzed. The prominence of W-REM and transition-related features like W-N1-REM suggests that muscle tone regulation becomes disconnected from cortical state control during the vulnerable sleep-wake boundary periods that characterize sleep paralysis episodes in NT1.

**Figure 4.**
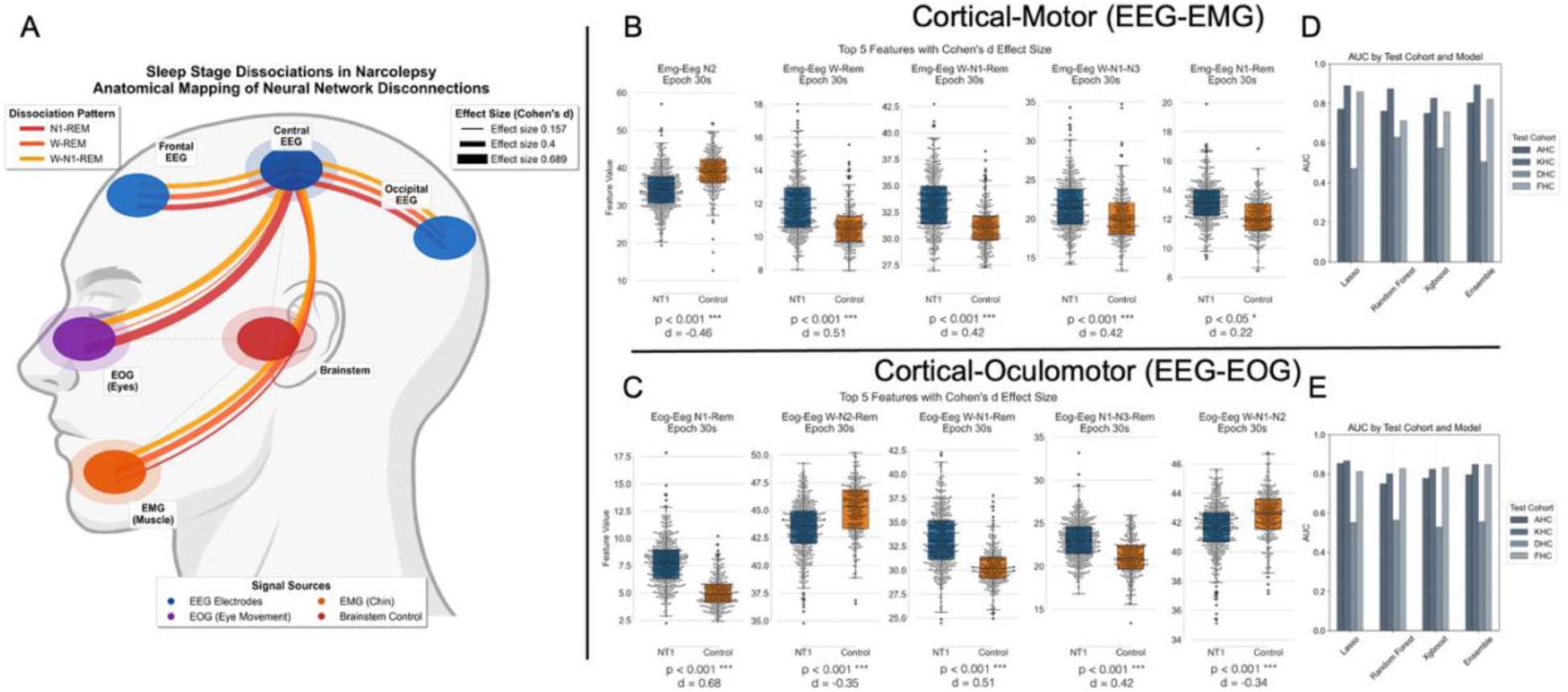
Cross-Modal Sleep Stage Dissociations in Narcolepsy Type 1: System-Wide Network Disconnections. **A)** Anatomical mapping of sleep stage dissociations showing hierarchical disconnection patterns across cortical-oculomotor (EOG-EEG), cortical-motor (EEG-EMG), and inter-cortical (C3-C4) networks. Line thickness represents effect size magnitude (Cohen’s d), with strongest effects observed in cortical-oculomotor connections (d=0,689), followed by inter-cortical (d=0.611) and cortical-motor (d=0.512) dissociations. Colors indicate dissociation patterns: N1-REM (red), W-REM (orange), and W-N1-REM (yellow) stage mixing. **B)** Top EEG-EMG dissociation features showing cortical-motor disconnection patterns. NT1 patients demonstrate elevated dissociation across multiple stage-mixing combinations, particularly during wake-sleep transitions. **C)** Top EEG-EOG dissociation features comparing NT1 patients (blue, N=452) and controls (orange, N=271). All features show significantly higher dissociation in NT1 (p<0.001) with large effect sizes. **D)** Leave-one-cohort-out cross-validation performance for EEG-EOG features across four cohorts, showing robust diagnostic utility (ensemble AUC range: 0.606-0.946) with individual algorithms include LASSO, Random Forest (RF), XGBoost (XGB), and ensemble methods. **E)** LOCO validation performance for EEG-EMG features, demonstrating consistent but more moderate diagnostic accuracy compared to cortical-oculomotor dissociations.

#### EEG-EOG Cortico-Oculomotor Disconnections

EEG-EOG dissociations demonstrated large effect sizes, with the strongest discriminator being EOG-EEG N1-REM stage mixing (Cohen’s d = 0.69, OR = 6.16, p < 0.001). Multiple EEG-EOG features showed large effect sizes, like N1-N3-REM mixing (d = 0.42, OR = 3.25, p < 0.001), indicating profound mixing disconnections between cortical sleep states and oculomotor control in NT1 (Fig. 4C). The magnitude of these oculomotor dissociations was remarkable, with NT1 patients showing up to 6-fold higher likelihood of exhibiting elevated EEG-EOG stage mixing compared to controls. For the strongest feature (N1-REM), NT1 patients demonstrated mean dissociation values of 7.787 ± 2.067 compared to 5.068 ± 1.421 in controls, a 54% increase in cortical-oculomotor disconnections. Like regional brain EEG electrode stage mixing dissociations, the most significant features involved combinations of sleep stages involving REM sleep with 4 of the 5 most significant features involving REM sleep.

Using the same dataset as regional EEG analysis (N=147 NT1 patients, N=168 controls), we evaluated both cortical-motor (EEG-EMG) and cortical-oculomotor (EEG-EOG) dissociation features through leave-one-cohort-out cross-validations. Cortical-oculomotor features demonstrated superior diagnostic performance (ensemble AUC = 0.800 ± 0.136, AP = 0.774 ± 0.175, Fig. 4E) compared to cortical-motor patterns (ensemble AUC = 0.779 ± 0.153, AP = 0.762 ± 0.184, Fig. 4D). Both cross-modal analyses showed between-site heterogeneity, with cortical-oculomotor features being most robustness across clinical sites. The KHC cohort achieved high performance for both modalities (EEG-EOG ensemble: AUC = 0.975; EEG-EMG ensemble: AUC = 0.919), with cortical-oculomotor features reaching high precision (AP = 0.992 vs. 0.976). The FHC cohort maintained strong discrimination for both analyses (EEG-EOG: AUC = 0.817; EEG-EMG: AUC = 0.830), while the AHC cohort showed comparable performance (EEG-EOG: AUC = 0.778; EEG-EMG: AUC = 0.807). The challenging DHC cohort also demonstrated better performance for cortical-oculomotor features (AUC = 0.630) than cortical-motor patterns (AUC = 0.558). Cortical-oculomotor models exhibited higher-weighted discriminative features compared to cortical-motor patterns. EOG-EEG N1-REM mixing emerged again as the dominant biomarker (coefficient=2.855, 100% occurrence, d=0.63, *p*_*fdr*_=7.0e-14), showing four-fold higher importance than the strongest cortical-motor feature (EMG-EEG N3-REM: coefficient=0.777, d=-0.03, *p*_*fdr*_=0.89). Complex multi-stage REM combinations dominated cortical-oculomotor selection, with N1-N3-REM and W-N2-N3-REM mixing both achieving 100% occurrence and high coefficients (2.734 and 1.306 respectively).

Cortical-motor models showed preference for different pathological patterns, emphasizing NREM stage transitions with N2-N3 mixing and N1-N2-REM combinations as the most consistently selected features (both 100% occurrence, coefficients ∼0.24). Wake-transition instability was prominent in both modalities but manifested differently: cortical-oculomotor features emphasized W-N1-REM combinations relating to hypnagogic hallucinations, while cortical-motor features highlighted W-N1-N2 patterns associated with sleep paralysis vulnerability. The markedly superior performance and higher feature weights of cortical-oculomotor dissociations suggest that eye movement control represents the most vulnerable system to sleep-wake state mixing in NT1.

### Integrated Multi-Modal Model

To evaluate whether combining regional EEG patterns with cross-modal dissociations enhances diagnostic accuracy beyond individual modalities, we used a multi-modal approach incorporating all 273 features across cortical (regional EEG), cortical-motor (EEG-EMG), and cortical-oculomotor (EEG-EOG) systems. This analysis employed the same leave-one-cohort-out cross-validation framework to ensure fair comparison with individual modality performance. The integrated model demonstrated robust diagnostic performance (ensemble AUC = 0.784 ± 0.119, AP = 0.788 ± 0.16), similar to individual modalities (Fig. 5A). Individual algorithm performance remained consistent with single-modality analyses, with Random Forest achieving the highest individual performance (AUC = 0.784±0.129, AP=0.763±0.191). The convergent performance across different modalities indicates that NT1 creates system-wide sleep dissociations rather than isolated regional dysfunction. Feature importance analysis revealed distinct algorithmic pathways to NT1 diagnosis. LASSO regression demonstrated strong preference for cortical-motor dissociations (normalized importance: 0.60), emphasizing high-magnitude EMG-EEG features. In contrast, Random Forest and XGBoost employed more balanced strategies, with comparable weighting across cortical (0.25-0.31) and cortical-oculomotor (0.21-0.30) systems (Fig. 5B). This algorithmic diversity achieving similar diagnostic performance strengthens confidence in the robustness of sleep-wake dissociation biomarkers across multiple analytical approaches.

**Figure 5.**
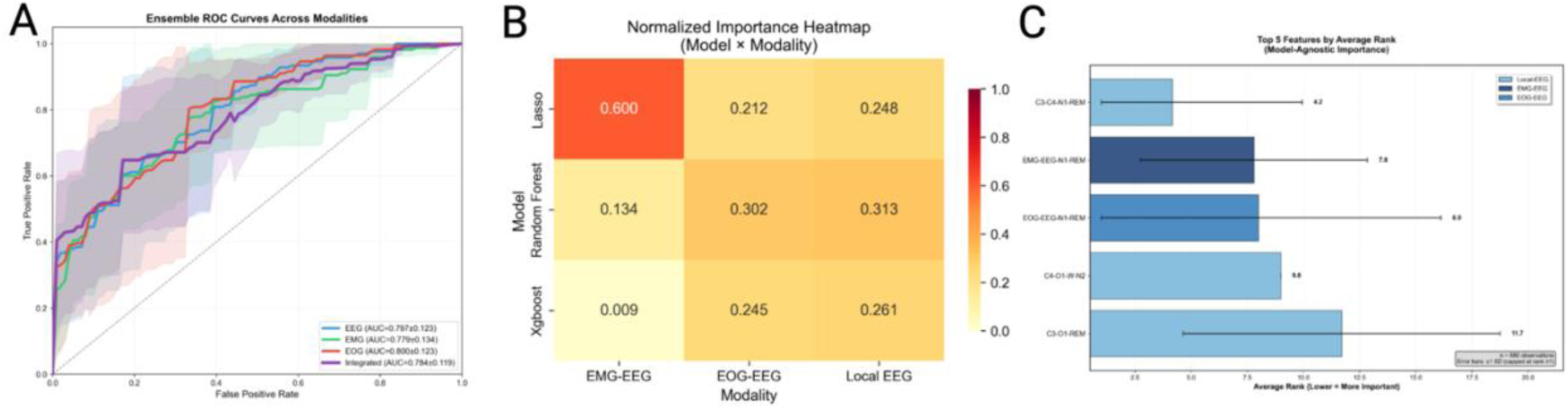
Integrated Multi-Modal Sleep Stage Dissociation Analysis Confirms System-Wide NT1 Pathophysiology. A) Ensemble ROC curves demonstrate similar diagnostic performance across neural systems, with confidence bands showing variation across leave-one-cohort-out validation folds. B) Algorithm-specific modality weighting reveals distinct pathways to NT1 diagnosis: LASSO emphasizes cortical-motor dissociations, while Random Forest and XGBoost favor balanced cortical and cortical-oculomotor patterns. C) Cross-algorithm consensus ranking identifies universal NT1 biomarkers, with N1-REM stage mixing emerging as the dominant pathological signature across cortical (C3-C4), motor (EMG-EEG), and oculomotor (EOG-EEG) systems. Error bars indicate inter-algorithm variability; smaller bars reflect stronger consensus. All analyses used leave-one-cohort-out cross-validation across four clinical sites (N=147 NT1 patients, N=168 controls).

### N1-REM Dissociation Emerges as Core Biomarkers Across All Modalities

The integrated analysis identified remarkably consistent core biomarkers that transcend individual neural systems. N1-REM stage mixing emerged as the universal pathological signature (Fig. 5C), appearing as the top feature in all three modalities: C3-C4 N1-REM (regional EEG d=0.61, *p*_*fdr*_=3.8e-11), EMG-EEG N1-REM (cortical-motor, d=0.22, *p*_*fdr*_=0.03), and EOG-EEG N1-REM (cortical-oculomotor d=0.69, *p*_*fdr*_=3.6e-16). This convergent pattern across independent neural systems validates N1-REM dissociation as a fundamental mechanism underlying NT1 pathophysiology. The C4-O1 electrode pair demonstrated vulnerability, ranking among top features across multiple algorithms (C4-O1 N1-REM mean importance: 2.73 ± 3.24 in LASSO models, d=0.46, *p*_*fdr*_=1e-4), suggesting that right central to left occipital connectivity represents a critical pathway for sleep-wake state coordination.

### Coherence analysis across sleep stages reveal modest changes in NT1

Coherence is a mathematical technique (see methods) that quantifies the frequency and amplitude of the synchronicity of neuronal patterns of oscillating brain activity. This technique quantifies synchronicity measured between spatially separated scalp electrodes. Brain regions represented by central (C3, C4) and occipital (O1, O2) electrode pairs across all cohorts (n=147 NT1 patients and n=168 controls), with additional frontal (F1, F2) electrode analysis in a subset of participants (n=40 NT1, n=39 controls), gathered across 5 different locations across the world (Austria, Denmark, France, Italy, Korea) were used^39,40^ (Supplementary table 1. As shown in supplementary Figure 1, panel A, coherence (adjusted by sex, age, BMI and cohort) was decreased in frontal/central areas and increased in occipital regions across all sleep stages and power bands (note that sample size was significantly lower for derivations involving frontal electrodes). Interestingly, however, the effect was stronger in C3-C4 and achieved statistical significance after False Discovery Rate (FDR) correction only in REM sleep in the delta range (supplementary Figure 1, panel A). These results suggest a minor decrease in coherence, with anterior dominance, more pronounced during REM sleep at low frequency.

### Regional Vulnerability Patterns correlate with CSF Hypocretin-1 in patients with low levels

Although hypocretin-1 is often undetectable in the CSF, many patients exhibit partially detectable levels, indicating remaining orexin activity in some subjects. We thus next explored how CSF hypocretin-1 levels, which may reflect the degree of orexin deficiency with NT1, affect global (EMG-EOG-EEG) and local EEG lead dissociations. In these analyses, the left central-to-occipital pathway (C3-O1) demonstrate the strongest effects, C3-O1_W-REM (r=-0.41, p=8.80e-08) and C3-O1_W-N3-REM (r=-0.40, p=1.52e-07), with the left frontal-central circuit (C3-F3_W) showing the strongest correlation (r=-0.50, p=2.23e-05), validating left hemisphere vulnerability observed in Figure 2B. Cross-modal dissociations revealed the following hierarchy: local EEG circuits (r=-0.40 to −0.50) > cortical-motor networks and cortical-oculomotor systems (EMG-EEG_W-N1-REM: r=-0.35, p=7.58e-09; EOG-EEG_W-N1-REM: r=-0.35, p=1.37e-08). Although higher levels of dissociation were associated with lower CSF hypocretin-1 levels across all modalities, the fact that strongest correlations were observed at the level of the EEG rather than globally suggest that the loss of orexin may first affect local cortical circuits. Interestingly, pure N2 as well as W-N1 dissociation shows the opposite relationship (Fig. 6A) consistent with findings showing opposite effects for N2, as observed notably in oculomotor-dissociation.

**Figure 6.**
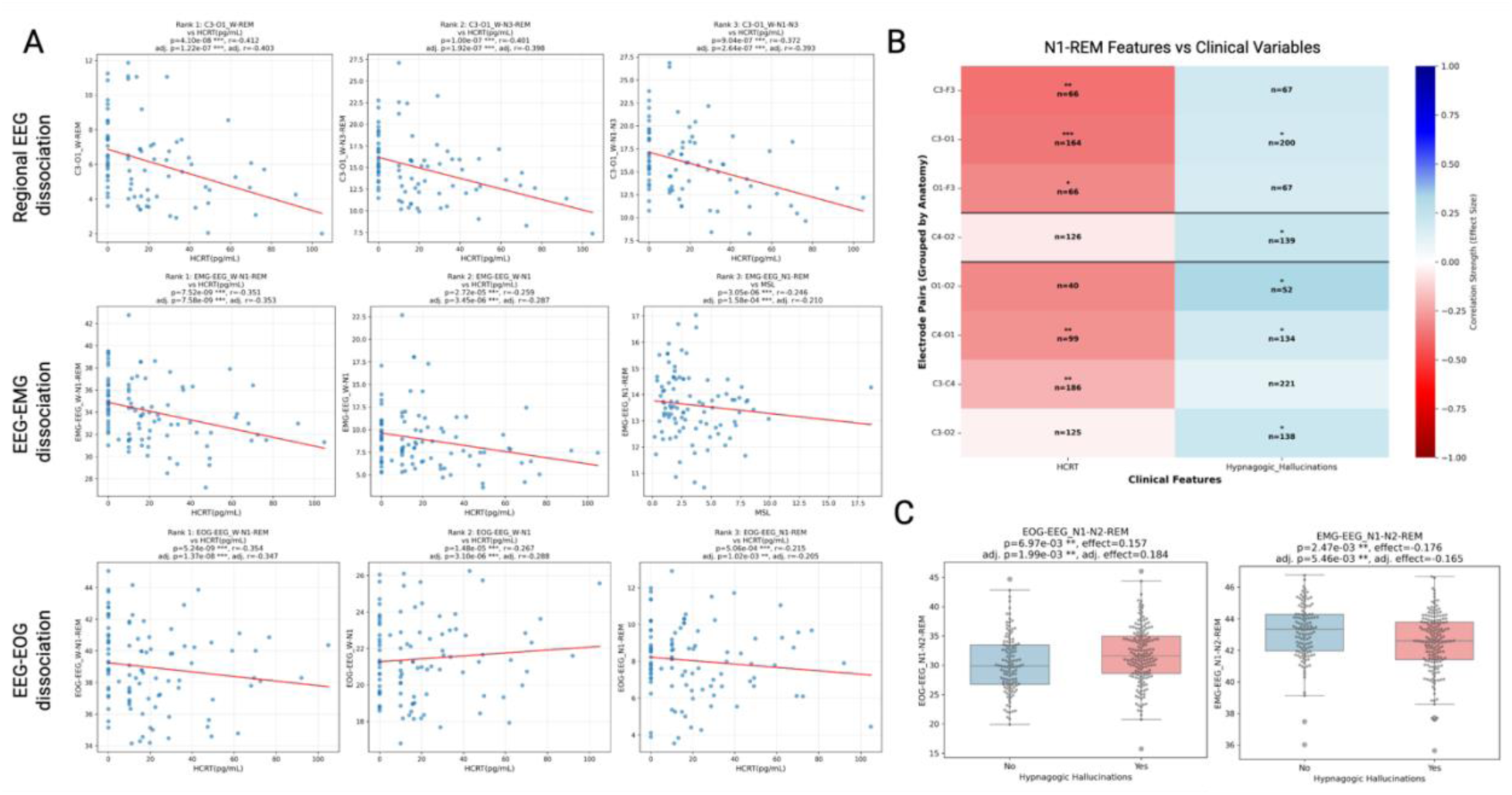
Clinical Correlations Reveal System-Specific Vulnerability and Opposing Symptom Mechanisms in Narcolepsy Type 1. A) Scatter plots show the top three correlations for each modality. Local EEG circuits demonstrate the strongest vulnerability to orexin deficiency, followed by cortical-motor networks, and cortical-oculomotor systems. B) Regional anatomical specificity of N1-REM stage mixing correlations with clinical features. Heatmap displays correlation coefficients for electrode pairs (rows) versus CSF hypocretin-1 and hypnagogic hallucinations (columns). Left hemisphere circuits show dominant vulnerability to orexin deficiency (C3-F3, C3-O1, C3-C4 showing strongest negative correlations with HCRT), while right-sided occipital circuits show slightly stronger correlation with hypnagogic hallucination frequency. Significance levels: *p<0.05, **p<0.01, ***p<0.001. C) Opposing symptom mechanisms across neural systems. Box plots compare N1-N2-REM stage mixing dissociation in patients with and without hypnagogic hallucinations (N=180 with vs 154 without). Cortical-oculomotor dissociation (EOG-EEG) facilitates hallucinations with higher dissociation in symptomatic patients (r=+0.18, p=5.46×10⁻³). while cortical-motor dissociation (EMG-EEG) shows protective effects with lower dissociation in symptomatic patients (r=-0.17, p=5.46×10⁻³).

### Symptomatic correlations of EEG, EOG and EMG stage mixing dissociations

Stage mixing differences across cortical area and between EEG, EOG and EMG are expected to reflect key aspects of narcolepsy symptomatology. To test this, we examined associations between self-reported sleep paralysis or hypnagogic hallucinations (presence or absence) and the occurrence of stage mixing across modalities (Figure 6). Although only the presence or absence of symptoms was available at the time of PSG, significant associations emerged between hallucination and ocular-cortical dissociations involving REM: EOG-EEG_N1-N2-REM (r=+0.18, p=1.99e-03), EOG-EEG_N1-N2 (r=+0.17, p=5.55e-03), and EOG-EEG_W-N1-REM (r=+0.16, p=8.30e-03). In contrast, cortical-motor dissociations showed inverse relationships: EMG-EEG_W-N1-N2-N3-REM (r=-0.18, p=1.96e-03) and EMG-EEG_W-N2-REM (r=-0.17, p=3.84e-03). These results suggest that hallucinations in NT1 are more strongly associated with episodes in which phasic REM-like ocular activity occurs while cortical signals remain in lighter or mixed states, a form of incomplete or spatially dissociated REM activation. (N1-N2-REM in Figure 6C)

At the regional EEG analysis level, correlations for hallucinations were consistent with a central-occipital dominance as shown in Figure 2A, consistent with involvement of the visual cortex. Right-sided occipital circuits correlated most with hypnagogic hallucinations: C4-O2_W-N2 (r=+0.27, p=1.70e-03) and C3-O2_N1-N2 (r=+0.27, p=1.73e-03), while bilateral occipital disconnection (O1-O2_W-REM) correlated with SOREMP frequency (r=-0.35, p=1.49e-03).

Sleep paralysis showed significant correlations only with regional EEG dissociation patterns, particularly right-sided central-occipital circuits (C4-O2_N1-N2-N3-REM: r=+0.23, p=2.48e-03) and occipital-frontal pathways (O1-F3_N1: r=+0.42, p=2.88e-03), while cortical-motor and cortical-oculomotor dissociations showed no significant associations at p<0.01.

Dissociation features correlated robustly with established NT1 diagnostic measures, confirming clinical relevance. Specifically, EMG-EEG_N1-REM correlated with mean sleep latency (r=-0.21, p=1.58e-04). Because MSLT and SOREMP data were available only for NT1 patients, these correlations were interpreted within this group and not as direct comparisons with controls.

## Discussion

In narcolepsy, we found increased coherence in occipital regions during wake, and less preeminently across all sleep stages, and relatively reduced frontal coherence during REM sleep only. Our study shows changes in coherence across sleep stages that reflect exacerbated physiological sleep stage specific changes in orexin deficient narcolepsy (NT1). Indeed, studies in the medical literature demonstrate that during wakefulness, high frequency-band coherence is generally increased across cortical regions, including the occipital cortex, reflecting integrated cortical processing. In contrast, during REM sleep, there is a marked reduction in gamma coherence, particularly in the frontal cortex, with some preservation or even relative increase in posterior (occipital and temporoparietal) regions, especially during phasic REM microstates.

Prior work has shown that nocturnal sleep in NT1 is characterized by sleep stage sequencing abnormalities, notably fast transitions from wake to REM sleep that bypass the classical 90-minute sleep cycle, reduced circadian gating of REM sleep outside of the early morning zone, and absence of typical occurrence of a small period of N2 sleep prior to a transition to REM sleep^38,44–47^. Additionally, increased N1 sleep and sleep stage transitions are observed as typical abnormalities following manual sleep scoring^48–50^. This increase was later found to frequently consist of a mix of N1, wake, REM sleep features reflecting the occurrence of a new, mixed sleep stage. This stage is easily observable through the analysis of sleep stage probability distributions when a neural network trained on normal sleep is applied to NT1 sleep^39^.

Use of deep learning predictions of sleep stages, including mixed probability, combined with HLA has been found to be highly predictive of an NT1 diagnosis^39,42,51^. As these features were obtained through the analysis of global PSG signals, we next wondered if mixed states, as obtained using predictions of sleep stages using entire PSG montages (EOG, EMG, and multiple EEG derivation, at minima central and occipital), are reflecting differential sleep stage occurrences across different regions of the brain or a global mixing event. To test this, we trained neural networks on each derivation individually using U-sleep to predict manually scored sleep stages and compared probabilities of sleep stages and occurrence of mixed states across derivations. U-sleep uses the U-Net architecture and can process PSGs at various resolutions without explicit modeling of temporal dependencies between adjacent epochs, unlike Long Short-Term Memory (LSTM) models. On global PSGs, U-Sleep demonstrates robust performance, with overall accuracy and Cohen’s kappa values that are within the range of inter-scorer variability seen among human experts^52^. To extend on this model, we also trained an EEG and EOG only network. Conceptually, symptoms such as sleep paralysis may reflect a global EEG pattern resembling wakefulness combined with EMG atonia typical of REM sleep. Conversely, REM Behavior Disorder (RBD) represents the opposite pattern, with REM-like EEG activity but loss of atonia.

We found substantial differences in the occurrence of sleep mixing, most notably between REM sleep, N1 sleep and wake, following a generally decreasing fronto-occipital gradient. This surprising result indicates that sleep stage mixing, as defined by prior deep learning model, can manifest more prominently in certain cortical regions than in others. Interestingly, differences in sleep stage mixing across cortical electrodes were not systematically directional (aka no single region consistently showed higher levels across the night). Instead, mixing fluctuated across electrodes, with a temporal dynamic indicating that short epochs of discordance lasting a few minutes were most discriminative of NT1 compared to controls, particularly in central regions. Although these intervals appear relatively long compared to self-reported durations of hallucinations or sleep paralysis, typically described as lasting less than 2min^30^, such estimates are based on subjective recollection and are prone to significant bias. Therefore, these physiological durations should not be directly equated with patient-reported experiences, which may underestimate or exaggerate their true length. Consistent with this interpretation, a recent study on lucid dreaming in narcolepsy found that lucid episodes were characterized by longer REM sleep periods (11-14 min vs 8 min)^53^, suggesting that prolonged REM stability may underlie more vivid or conscious experiences during sleep.

Hypodensity probabilities of sleep stages are without a doubt a simplification of the complexity of EEG patterns occurring during sleep. In this context, the occurrence of a mixed state pattern must be viewed in the context of recent high density EEG work^54^ or intracranial EEG recordings^26,55^ during REM sleep, some of it likely corelative to dream content, its remembrance, and/or perhaps the associated muscle paralysis. Intracranial recordings have further shown that phasic and tonic REM substates exhibit distinct regional spectral profiles, with occipital-temporal and medial frontal regions displaying opposite power gradients, supporting focal sleep regulation within REM states^56^. Of particular interest has been the description of slow wave in REM sleep^57^, primarily located in primary sensory and motor areas, possibly reflecting decreased activity in these areas most immediately connected to sensory input and locomotor output^58^. (EEG slow frequencies between 1 and 4 Hz during sleep are associated with neuronal down-states and bi-stability^59^, which prevent the emergence of stable causal interactions among cortical areas^60^, and have been linked to the loss of consciousness^61^. Slow wave sleep activity, together with theta, is intrinsic to saw tooth waves (but not exclusively^57^) and has been shown to also be time locked with fast gamma activity during REM sleep^26^. Although this is not formally testable in our low-resolution EEG montage, increased mixing was more preeminent in central and frontal areas, and notably in the frontal areas. This could correlate with increased awareness during dreaming experiences in narcolepsy as reported by others in normal dreamers^62^. Another candidate unexplored in this study would be the so called parieto-occipital “hot zone” where decreased slow wave activity in both REM and non-REM sleep has been correlated with dream recall^63^. Intriguingly, although we found limited correlations between occurrence of symptoms and local stage mixing (beside severity measures), presence of hypnagogic hallucinations correlated with EEG mixing derivations involving occipital areas (for example C3-O1 or C4-O2), or EEG-EOG dissociations, notably on the left (dominant) side (Figure 6). Perhaps stage mixing in occipital area reflects visual aspects of dreaming^64^, which are frequent in the conscious experience of hypnagogic hallucinations.

More disappointingly, we found no correlations between self-reports of sleep paralysis in NT1 and dissociative combinations involving EMG. Our initial hypothesis was that dissociated predictions of REM sleep versus wake using EEG versus EMG would have been most powerful discriminant for such symptoms. Such abnormalities are strong predictor features in the differentiation of wild type versus mouse with orexin deficiency^65^. This absence of association may reflect the fact most adult patients with narcolepsy report sleep paralysis, whereas such reports are rare in prepubertal children ^66–68^, adding a maturational aspect to the issue. Additionally, most patients in this sample reported sleep paralysis, and no information on symptoms frequency were included, making it hard to study. Finally, we did not have reports of these symptoms specifically for the night being analyzed here, thus reducing our ability to truly explore how these symptoms manifest.

The study has strengths and many weaknesses. One strength is the study of multiple cohorts of NT1 and controls across multiple centers, with consistency of the results obtained, and the relative ease of interpretation of hypnodensity results. It is also likely that addition of local hypnodensity features (variation across electrodes) could further improve the diagnostic value of PSG recordings for the diagnosis of narcolepsy, as suggested in Figure 5. A major weakness, beside its conceptual nature reflected by sleep stage probabilities rather than the study of actual EEG or subcortical activity, is the fact a low-density EEG montage was used. As shown in our study and discussed by many others, aspects of sleep, whether NREM or REM sleep, are clearly local, and the resolution of a simple PSG montage is clearly insufficient for proper source localization. Further, as hinted by our data, local alterations are likely correlated, in the case of REM sleep, with the complexity of a sleep stage that includes multiple aspects: vivid dreaming, rapid eye movements, phasic and tonic muscle atonia, and absence of dream recollection, features that have likely neural network correlates that are disturbed in narcolepsy. It is thus likely that the careful study of a smaller number of patients using high density EEG in correlation with subjective reports of experiences when being woken up, could establish causality between symptoms and local pattern obtained. This, in turn, could inform us on the wide range of experiences individuals, narcoleptic or not, can experience during sleep, as parasomnia including those involving REM sleep are much more frequent than narcolepsy and commonly associated with psychiatric comorbidities and poor sleep hygiene^69,70^. This could also apply to the special case of REM behavior disorder, a common precursor of alpha-synucleinopaties notably Parkison disease and Lewy Body dementia^71^. Exploring response to therapy and further studying these dissociations across the normal population could reveal novel insights. We anticipate that studying local sleep mixing probabilities in other parasomnias, such as sleep waking, talking or night terrors, may reveal similar anatomical dissociations but in NREM-wake probabilities, with implication for our understanding of mental health and healthy sleep.

## Method

### Data used in the study

We utilized a multi-center dataset comprising polysomnographic recordings from five international hypersomnia cohorts (Supplementary Table 1): Korean Hypersomnia Cohort (KHC), Austrian Hypersomnia Cohort (AHC), Italian Hypersomnia Cohort (IHC), Danish Hypersomnia Cohort (DHC), and French Hypersomnia Cohort (FHC), as previously described in Stephansen et al.^39^ and further analyzed in Olsen et al.^40^. Note that these manuscripts also include data from China, but these were not included in this study due to the extreme age imbalance of cases versus controls, most Chinese cases being young children. For this analysis, we included only untreated NT1 patients and healthy controls, excluding other hypersomnia disorders to ensure diagnostic specificity. The age ranges for each cohort were as follows: AHC (NT1: 18-59 years, controls: 19-55 years), DHC (NT1: 13-61 years, controls: 23-50 years), FHC (NT1: 14-70 years, controls: 20-71 years), IHC (NT1: 9-67 years, controls: 11-53 years), KHC (NT1: 16-52 years, controls: 16-70 years). NT1 patients were diagnosed based on established criteria: documented cataplexy, positive Multiple Sleep Latency Test (mean sleep latency <8 minutes with ≥2 sleep-onset REM periods), and/or cerebrospinal fluid hypocretin-1 levels ≤110 pg/ml. Controls consisted of healthy subjects without sleep complaints or other hypersomnia symptoms. The multi-center design provides diverse recording environments with different equipment, patient populations, and clinical practices, enabling robust assessment of diagnostic generalizability across real-world clinical settings. For regional EEG analysis, we utilized the subset of recordings with the necessary electrode coverage (N=147 NT1 patients, N=168 controls). For cross-modal analysis, we employed the complete dataset with standard EMG and EOG channels available across all sites (N=452 NT1 patients, N=271 controls). A subset of participants had clinical symptom assessments including presence or absence of sleep paralysis, and hypnagogic hallucinations as a reported symptom (N=180 with and 154 without, no information on dream enactment behavior was available) and hypocretin measurements, which were used for clinical correlation analyses (N=292 with 89% NT1).

### Coherence analysis

We computed magnitude-squared coherence between EEG channel pairs to quantify frequency-specific coupling across cortical regions. Prior to spectral estimation, signals were resampled to 128 Hz, notch-filtered at mains frequency (50/60 Hz), high-pass filtered at 0.5 Hz, and re-referenced to linked mastoids (A1/A2) to ensure a consistent reference across cohorts. For each PSG, 30-s epochs were grouped by AASM stage (W, N1, N2, N3, REM). Within each stage, we estimated time-frequency coherence *C*_*xy*_(*f*, *t*) from all available electrode pairs using sliding 5-s windows with 1-s overlap. Cross- and auto-spectral densities were obtained via multitaper estimation, and coherence was defined as the normalized cross-spectrum,

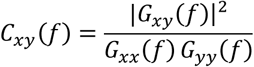

Coherence was evaluated from 0.2–30 Hz at 0.2-Hz resolution. For each stage, we averaged *C*_*xy*_(*f*, *t*) over time and then within canonical bands to obtain band-limited coherence per pair (δ: 1–3 Hz; θ: 4–7 Hz; α: 8–12 Hz; β: 13–30 Hz). In addition to these stage-specific features, we also extracted quarter-of-night and whole-night features. The primary analysis used central/occipital leads (C3, C4, O1, O2), yielding 6 pairs and 6 pairs × 4 bands × (5 stages + 4 quarters + 1 whole) = 240 features. In a frontal-extended subset (F3, F4 present), we analyzed 15 pairs for a total of 15 × 4 × 10 = 600 features. Code to reproduce these steps is available in our psg-coherence repository.

### Staging Model

To investigate regional sleep stage dynamics, we utilized electroencephalographic (EEG) data from multiple scalp locations following the standard 10-20 system. As illustrated in Figure 1A, we focused on key electrode positions including frontal (F3, F4), central (C3, C4), and occipital (O1, O2) derivations, allowing for comprehensive cortical coverage. All EEG derivations were re-referenced to the contralateral mastoid (C3–M2, C4–M1, O1–M2, O2–M1, F3–M2, F4–M1) following AASM standards. For sleep stage classification, we used the U-Sleep-EEG v2.0 neural network (Figure 1B), which transforms raw EEG signals through sequential encoding and decoding blocks into probabilistic sleep stage distributions. This automated scoring system was trained on one EEG input channel of PSG recordings from 25,696 participants and achieves accuracy on par with the best human experts^43^. The network’s softmax output produces a probability density function over the five sleep stages (Wake, N1, N2, N3, REM), with probabilities summing to 1 for each electrode and time point. This probabilistic approach enables detection of subtle transitions and sleep stage mixing phenomena typically overlooked in traditional scoring. To produce EMG and EOG predictions we used a USleep model pre-trained on EEG, EOG and EMG channels and applied a two-stage transfer learning approach^72^ with a progressive unfreezing strategy to adapt the model for single-channel EOG and EMG sleep staging. For fine-tuning we used the Danish Center for Sleep Medicine (DCSM) dataset consisting of 255 randomly selected overnight lab-based PSG representing a diverse Danish cohort collected between 2015-2018^43^ and the Wisconsin Sleep Cohort (WSC), a longitudinal study of sleep disorders with overnight in-laboratory polysomnography^73,74^. Stage 1 involved training only the first convolutional layer and classifier head while keeping all other parameters frozen, using a learning rate of 1 × 10^−4^ for 10 epochs. Stage 2 consisted of full model fine-tuning with all parameters trainable at a reduced learning rate of 1 × 10^−5^ for up to 20 epochs. Early stopping with a patience of 10 epochs was applied during Stage 2 to prevent overfitting, monitoring validation loss for convergence. This progressive unfreezing strategy allows the model to first adapt its initial feature extraction to the target domain before fine-tuning the entire network architecture. During the fine-tuning we used a batch size of 64 training the model on a single gpu. The progress was monitored by validation loss and accuracy, also monitoring sleep stage specific accuracy. For EMG the model struggled with NREM stages and provided more accurate results for REM while the EOG model struggled with REM stage classification. The models were tested on a hold-out dataset based on the Inter-scorer Reliability Cohort (IS-RC), which consisted of 70 PSGs scored by 6 expert scorers across three locations in the United States^75^ (Supplementary Figure 2).

### Stage-mixing Dissociation Features

The stage-mixing features we developed capture simultaneous occurrence of different sleep stages across brain regions, representing the core pathophysiological signature of narcolepsy. This approach was inspired by Stephansen et al.^39^, which we adapted for pairwise electrode-specific hypnodensity as well as EEG-EMG and EEG-EOG comparisons where the average across electrodes is used. The formula below shows the stage-mixing feature for two electrodes and can be analogously applied to EEG-EMG and EEG-EOG comparisons. The stage-mixing feature Φ_*n*_(*S*_*k*_, *E*_*a*_, *E*_*b*_) quantifies the dissociation between electrode pairs by measuring the absolute difference in combined sleep stage probabilities:

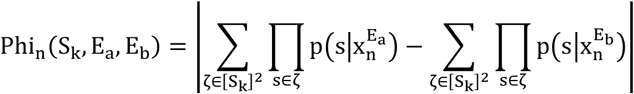

where *S*_*k*_ represents a set of *k* sleep stages (e.g., *k* = 2 for combinations like ‘W-N1’ or ‘N2-REM’), and [*S*_*k*_]^2^ denotes all possible stage combinations. To obtain the final stage-mixing feature, we average across the time series and apply a normalization factor 1/*k* to counter the multiplication effect:

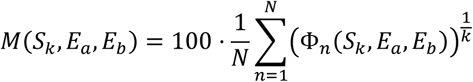

This approach generated 31 stage combinations across 6 electrode combinations, resulting in 186 proto-features that capture regional cortical dissociations. The same 31 stage combinations were applied to cross-modal comparisons, generating additional features for cortical-motor dissociations (EEG-EMG, comparing each EEG electrode to chin EMG) and cortical-oculomotor dissociations.

### Statistical Analysis

We analyzed coherence and dissociation features by first adjusting each feature for key covariates, then testing NT1-control differences on the adjusted values. Subjects with missing data in the outcome or any covariate were excluded from the corresponding analysis. Covariates included age, BMI, sex, study cohort, and mean sleep stage probabilities, with the latter included to control for known differences in overall sleep architecture. Adjustment was performed using ordinary least squares regression, with residuals carried forward for group comparison. NT1–control differences were assessed with two-sided Welch’s t-tests, and effect sizes were expressed as adjusted Cohen’s d (positive when NT1 > control). Multiple comparisons across all features were controlled using the Benjamini–Hochberg procedure, with significance defined as *p*_*FDR*_ < 0.05.

### Temporal Analysis

To determine the optimal temporal resolution for detecting regional dissociations, we analyzed variance ratios across multiple epoch lengths (1s, 3s, 15s, 30s) for all significant electrode pairs and features. The variance ratio serves as a measure of signal-to-noise ratio, quantifying how well each feature discriminates between NT1 patients and controls at different temporal scales:

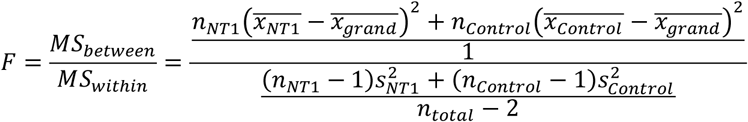

where 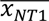 and 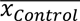 are group means, 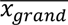 is the overall mean, and *s*^2^ denotes the group variances. Higher F-ratios indicate better discrimination between groups at that temporal resolution.

Sustained dissociation periods were identified by converting the N1-REM proto-features time series to a binary sequence using an amplitude threshold, then finding the longest continuous run of epochs above this threshold. The binary conversion process classified each epoch as "dissociated" if the feature value exceeded the threshold, or "synchronized" otherwise. For each subject, we extracted the duration of the longest continuous dissociation period within their recording. Subjects were classified as NT1 if their longest continuous dissociation period exceeded a duration threshold, with both amplitude and duration thresholds optimized using grid search cross-validation to maximize diagnostic accuracy. This approach tests whether the temporal persistence of dissociation, rather than just its magnitude or frequency, provides discriminative power for NT1 diagnosis.

For clinical correlations spearman rank correlation coefficients were calculated between each stage-mixing dissociation feature and clinical variable to assess monotonic relationships while avoiding assumptions about data distribution. To control for potential confounding effects, which could mediate differences in severity, partial Spearman correlations were computed controlling for age, sex and onset age of excessive daytime sleepiness. The partial correlation coefficient was calculated as:

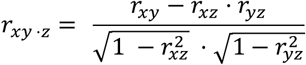

where *r*_*xy*⋅*z*_ is the partial correlation between feature *x* and clinical variable *y* controlling for age *z*, and *r*_*xy*_, *r*_*xz*_, and *r*_{*yz*} are the respective Spearman correlations. Statistical significance of partial correlations was assessed using t-statistics with degrees of freedom *df* = *n* − 2, where *n* is the sample size. Two-tailed p-values were calculated using the t-distribution.

### LOCO with Consistent Implementation Across Modalities

To evaluate the generalizability and clinical utility of sleep stage dissociation features, we employed a leave-one-cohort-out (LOCO) cross-validation strategy across all analyses^76^. This approach addresses the most clinically relevant scenario: whether diagnostic models trained at one set of sleep centers can accurately classify NT1 patients at previously unseen clinical sites with different recording equipment, patient populations, and scoring practices. The LOCO design iteratively trains models on three cohorts while testing on the fourth held-out cohort, repeated across all four combinations. This methodology prevents site-specific overfitting and tests true generalizability across diverse clinical environments, which is essential for real-world diagnostic deployment. Unlike traditional random cross-validation approaches that intermix patients from different sites, LOCO validation ensures that no information from the test site influences model training, providing the most stringent assessment of diagnostic robustness.

For individual modality analyses (regional EEG, cortical-motor EEG-EMG, cortical-oculomotor EEG-EOG), we applied the same LOCO framework using the dataset with complete electrode coverage for central occipital (N=147 NT1, N=168 controls) to enable direct performance comparisons. Each analysis employed identical machine learning pipelines including LASSO regression, Random Forest, XGBoost, and ensemble methods, with hyperparameters optimized within each training fold to prevent data leakage (Supplementary Table 4). For the integrated multi-modal analysis, we combined all 248 features (regional EEG dissociations, EEG-EMG stage mixing, EEG-EOG stage mixing) within the same LOCO framework, allowing algorithms to discover cross-modal feature interactions while maintaining the same validation standards. This approach tests whether combining information across cortical, motor, and oculomotor systems improves diagnostic accuracy beyond individual modalities, while ensuring that performance gains reflect genuine biological signal rather than methodological artifacts. The consistent LOCO methodology across all analyses enables fair comparison of diagnostic utility between regional EEG patterns, cross-modal dissociations, and integrated multi-system biomarkers, ultimately determining the optimal feature combination for robust NT1 diagnosis across diverse clinical settings.

## Data Availability

The polysomnography and associated clinical data analyzed in this study are available through the National Sleep Research Resource (NSRR) repository, subject to standard data use agreements.

https://sleepdata.org/datasets/mnc

---

**Supplementary Figure 1.**
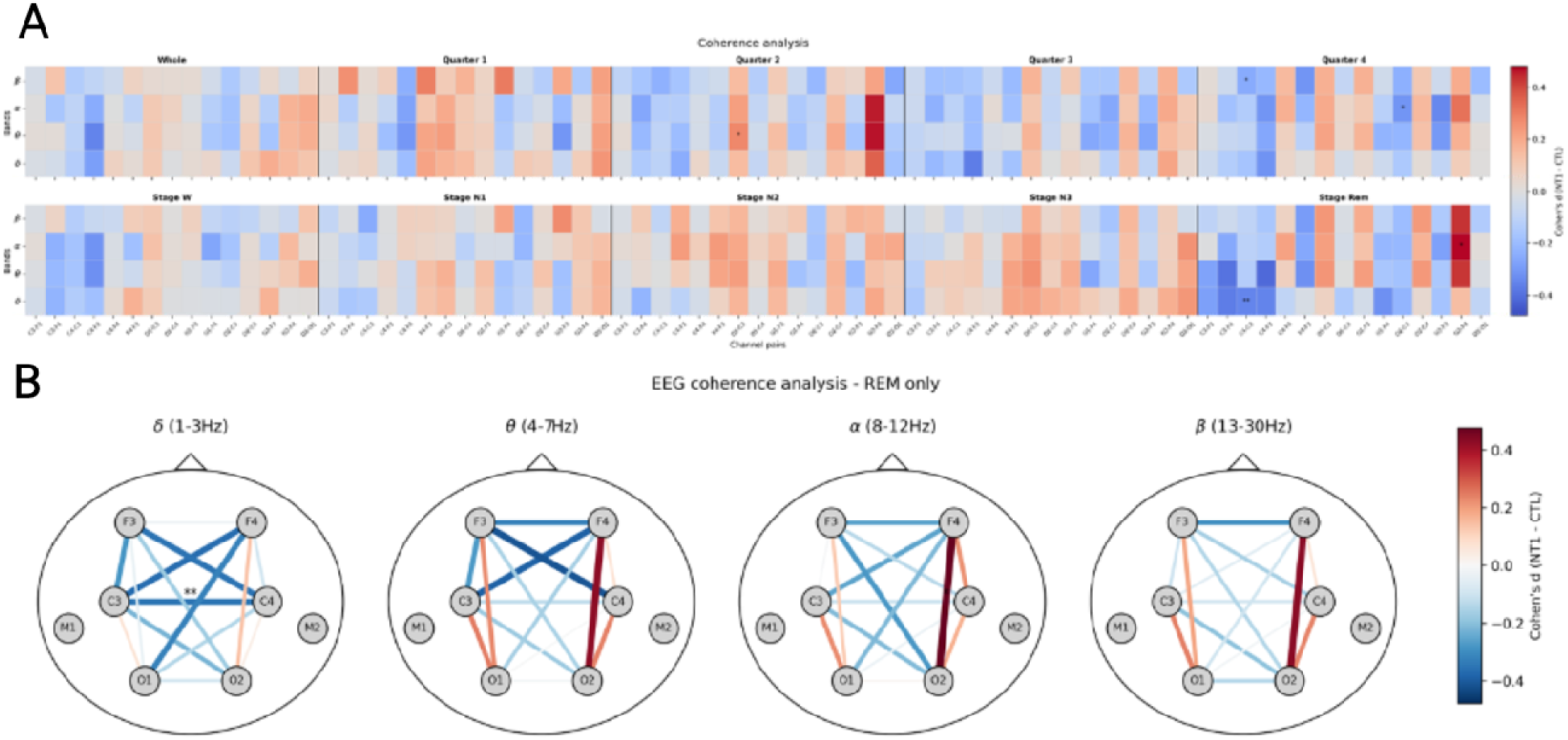
Coherence analysis. A) heat-map of Cohen’s *d* showing the effect size of coherence differences between narcolepsy type 1 patients and controls, organized by frequency band, sleep stage, and electrode scope. Positive values (red) indicate higher coherence in narcolepsy patients, while negative values (blue) indicate lower coherence compared to controls. After adjusting for key covariates, the only significant difference, corrected for a False Discovery rate of 0.05 is decreased interhemispheric (C3-C4) coherence in the low frequency delta range during REM sleep. (**=p<0.01). B). Similar trends are found in anterior and central but not occipital electrodes but decreased number of observations for frontal electrodes likely obscured statistical significance.

**Supplementary Figure 2.**
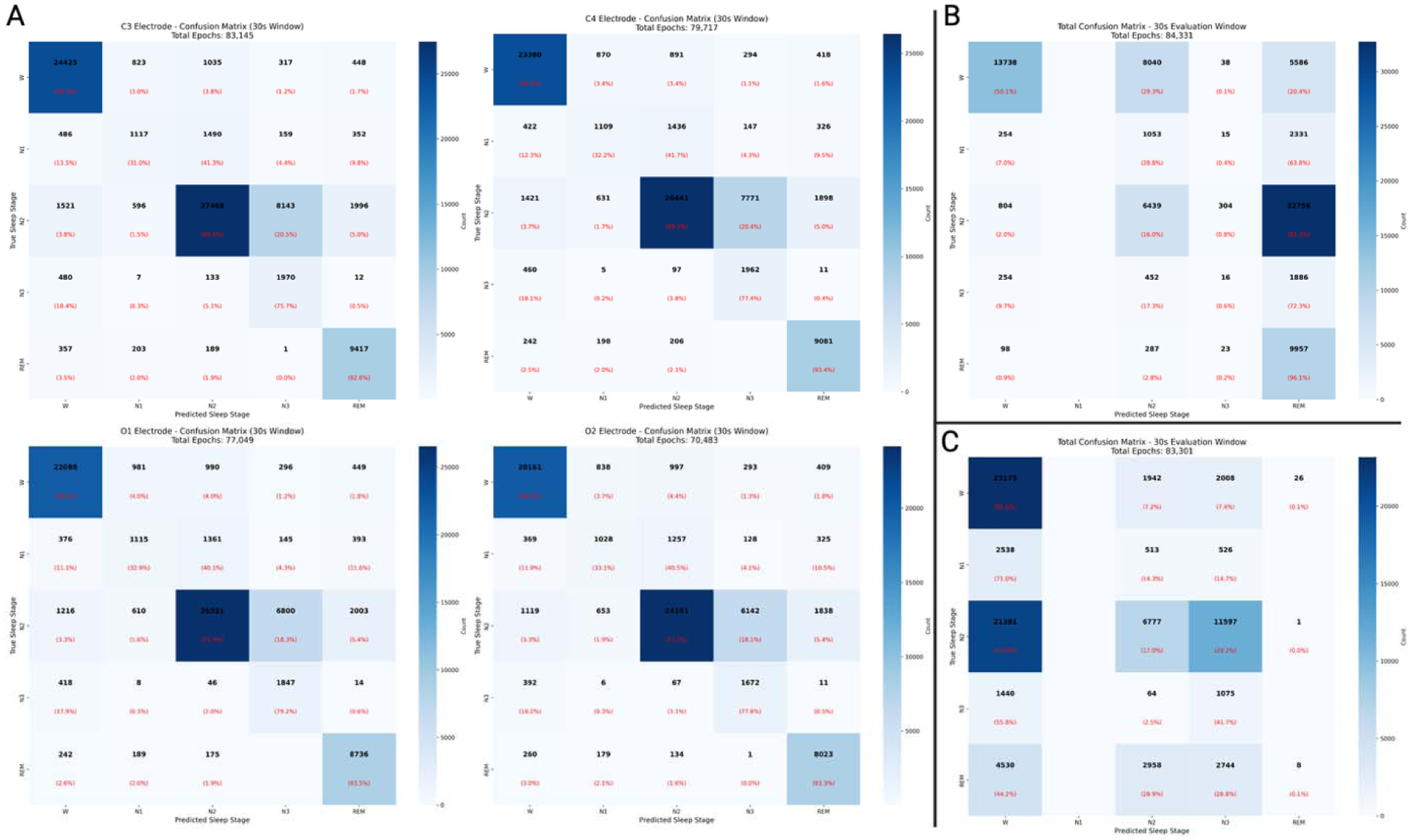
Electrode-level EEG, EOG, and EMG predictions of sleep stages. As described in the methods, we used different adaptations of the U-Sleep model using single-channel recordings from EEG (frontal, central, and occipital electrodes), EMG (submental), and EOG (horizontal eye movements) modalities. The predictions were evaluated on a holdout dataset as described in the methods. A) EEG-based models achieved the highest performance across all electrode locations, with consistent performance across electrode locations (accuracy ∼0.77-0.78, Cohen’s κ ∼0.67-0.68), with strong performance for Wake and REM (∼89-94%) but challenges in N1 classification (∼30-33%). B) EMG showed low performance but did show it distinguishes REM sleep due to muscle atonia detection. C) EOG demonstrated the poorest overall performance, especially for REM stage classification, consistent with the physiological reality that most REM sleep consists of tonic periods without eye movements that are indistinguishable from NREM sleep on EOG alone. These results confirm that EEG signals provide the most reliable features for automated sleep staging and narcolepsy-related sleep dissociation detection, while EOG and EMG offer complementary but limited information when used in isolation.

**Suppl table 1:** Cohort characteristics used in the study.

| Metric | Narcoleptic patients | Controls |
| --- | --- | --- |
| **Subjects** | 459 | 289 |
| **Age (mean±SD)** | 30 ± 17 | 35 ± 14 |
| **Sex (F)** | 42.7% | 53.7% |
| **BMI (mean±SD)** | 26 ± 5 | 24 ± 6 |
| **Nocturnal SOREMP ($\geq 2$)** | 96% (409/426) | 1.7% (2/117) |
| **with Cataplexy** | 96.5% (401/421) | 3.3% (5/142) |
| **with low CSF Hcrt ($<110$)** | 100.0% (347/347) | 0.0% (0/34) |
| **HLA positive** | 98.8% (421/426) | 15.7% (29/185) |
| **From Austria (AHC)** | 9.2% (42) | 39.4% (114) |
| **From Denmark (DHC)** | 4.6% (21) | 6.9% (20) |
| **From France (FHC)** | 41.2% (189) | 37.7% (109) |
| **From Italy (IHC)** | 35.7% (164) | 11.1% (32) |
| **From Korea (KHC)** | 9.4% (43) | 4.8% (14) |

**Supplementary Table 2.** validates our regional sleep stage dissociation findings against the established global diagnostic features identified by Stephansen et al. (2018). This comparison demonstrates that the same pathological stage combinations that distinguish narcolepsy type 1 patients at the global level (e.g., W-N2-REM, N1-REM) also manifest as regional dissociations across specific electrode pairs. Notably, while Stephansen’s top global feature was W-N2-REM mixing, our spatial analysis reveals C3-C4 N1-REM dissociation as the strongest regional discriminator. Stephansen’s original features were derived through diverse mathematical approaches including Shannon entropy, time-to-threshold measures, and prevalence calculations. Several Stephansen stage combinations including N1 (alone), N1-N2, and N3 (alone) did not achieve significance in our regional analysis above the adjusted p-value cutoff of p < 0.001.

| **Stephansen Stage Combination** | **Stephansen Selection Frequency** | **Top Regional Features** | **Effect Size Range (unadjusted Cohen's d)** | **Best p-value (adj.)** | **Electrode Pairs** |
| --- | --- | --- | --- | --- | --- |
| **W, N2, REM** | 1.00 | C3-C4_W-N2-REM | 0.604 | 1.83e-04 | C3-C4 |
| **W** | 0.82 | C3-O1_W, C3-O2_W | 0.456-0.506 | 5.32e-04 | C3-O1, C3-O2 |
| **REM** | 0.82 | C3-C4_REM, C3-O2_REM | 0.673-0.790 | 5.51e-09 | C3-C4, C3-O2 |
| **N2, REM** | 0.68 | C3-C4_N2-REM | 0.549 | 1.83e-04 | C3-C4 |
| **W, N2** | 0.68 | C3-O2_W-N2, C3-O1_W-N2, C4-O1_W-N2, C4-O2_W-N2, C3-C4_W-N2 | 0.469-0.532 | 3.41e-07 | C3-O2, C3-O1, C4-O1, C4-O2, C3-C4 |
| **W, N1** | 0.64 | C3-O1_W-N1, C3-C4_W-N1 | 0.452-0.488 | 1.83e-04 | C3-O1, C3-C4 |
| **W, N1, REM** | 0.55 | C3-C4_W-N1-REM | 0.644 | 9.63e-07 | C3-C4 |
| **W, N1, N3** | 0.55 | C3-O2_W-N1-N3, C3-O1_W-N1-N3, C4-O1_W-N1-N3, C3-C4_W-N1-N3 | 0.500-0.540 | 2.73e-06 | C3-O2, C3-O1, C4-O1, C3-C4 |
| **W, N1, N2, REM** | 0.55 | C3-C4_W-N1-N2-REM | 0.432 | 1.83e-04 | C3-C4 |
| **N1, REM** | 0.45 | **C3-C4_N1-REM**, C4-O1_N1-REM, C3-O1_N1-REM, C3-O2_N1-REM, C4-O2_N1-REM | **0.895-1.066** | **9.13e-11** | **C3-C4**, C4-O1, C3-O1, C3-O2, C4-O2 |

**Supplementary Table 3.** Top 20 features distinguishing NT1 from controls. Top 20 features ranked by FDR-adjusted p-values (Benjamini–Hochberg). Means are computed on covariate-adjusted residuals. Cohen’s d is positive when NT1 > Control. The full results for all features are provided in the Excel file “supplementary_features.xlsx”

| Feature's name | Mean NT1 | Mean CTL | Cohen's d | p | $p_{fdr}$ | NT1 | CTL |
| --- | --- | --- | --- | --- | --- | --- | --- |
| **EOG-EEG_N1-REM** | 0.334 | -0.559 | 0.689 | 3.14e-19 | 3.6e-16 | 446 | 267 |
| **C3-C4_N1-REM** | 0.321 | -0.513 | 0.611 | 6.66e-14 | 3.82e-11 | 373 | 233 |
| **C3-O2_W-REM** | 0.651 | -0.803 | 0.667 | 1.27e-13 | 4.86e-11 | 284 | 230 |
| **C4-O2_N1-REM** | 0.397 | -0.492 | 0.655 | 2.99e-13 | 8.56e-11 | 285 | 230 |
| **C3-O2_N1-REM** | 0.407 | -0.502 | 0.642 | 1.7e-12 | 3.89e-10 | 284 | 230 |
| **C3-C4_W-REM** | 0.451 | -0.721 | 0.579 | 3.53e-12 | 5.79e-10 | 373 | 233 |
| **C3-O2_W-N3-REM** | 1.24 | -1.53 | 0.626 | 3.35e-12 | 5.79e-10 | 284 | 230 |
| **EOG-EEG_W-N1-REM** | 0.399 | -0.667 | 0.515 | 4.78e-12 | 6.85e-10 | 446 | 267 |
| **C4-O2_W-REM** | 0.561 | -0.696 | 0.614 | 8.08e-12 | 1.03e-09 | 285 | 230 |
| **EMG-EEG_W-REM** | 0.284 | -0.474 | 0.513 | 1.17e-11 | 1.34e-09 | 446 | 267 |
| **C3-C4_W-N3-REM** | 0.843 | -1.35 | 0.57 | 2.05e-11 | 1.83e-09 | 373 | 233 |
| **C3-O2_W-N1-N3** | 0.826 | -1.02 | 0.602 | 2.08e-11 | 1.83e-09 | 284 | 230 |
| **C4-O2_W-N3-REM** | 1.13 | -1.39 | 0.598 | 1.97e-11 | 1.83e-09 | 285 | 230 |
| **C3-O2_W-N1** | 0.375 | -0.463 | 0.589 | 6.31e-11 | 5.17e-09 | 284 | 230 |
| **C4-O2_W-N1-N3** | 0.736 | -0.912 | 0.571 | 1.11e-10 | 8.5e-09 | 285 | 230 |
| **C4-O2_N1-N3-REM** | 0.799 | -0.99 | 0.568 | 1.54e-10 | 1.11e-08 | 285 | 230 |
| **C3-C4_N1-N3-REM** | 0.593 | -0.949 | 0.539 | 1.89e-10 | 1.27e-08 | 373 | 233 |
| **C3-C4_W-N1-REM** | 0.676 | -1.08 | 0.525 | 2.51e-10 | 1.6e-08 | 373 | 233 |
| **EMG-EEG_W-N2** | 0.434 | -0.724 | 0.475 | 4.41e-10 | 2.66e-08 | 446 | 267 |
| **C3-O2_N1-N3-REM** | 0.804 | -0.993 | 0.556 | 5.65e-10 | 3.24e-08 | 284 | 230 |

*Abbreviations: FDR, false discovery rate*

